# The association between medroxyprogesterone acetate exposure and cerebral meningioma among a Medicaid population

**DOI:** 10.1101/2025.06.26.25330350

**Authors:** Lindy M. Reynolds, Rebecca Arend, Russell L. Griffin

## Abstract

**BACKGROUND:** Medroxyprogesterone acetate (MPA) is a synthetic contraceptive that can be used orally or as a once-every-three-month injection (i.e., depot MPA [dMPA]). Prior research has reported an increased association between dMPA and cerebral meningioma, a cancer of the protective layers of the brain. These reported associations have been strongest among prolonged users (i.e., more than two continuous years), but prior research has been limited in generalizability to meningioma cases treated with surgery or cases derived from an administrative database of commercial insurance enrollees. The current study builds upon prior research by examining the association among public insurance enrollees utilizing both a non-active and active comparator.

**METHODS:** Utilizing Medicaid data, cases of cerebral meningioma were matched to up to ten controls based on age and year of Medicaid enrollment. A conditional logistic regression estimated odds ratios (ORs) and 95% confidence intervals (CIs) for the association between MPA and dMPA exposure and cerebral meningioma compared to both an active and non-active comparator.

**RESULTS:** Among 469 cases and 4690 matched controls, there was no association between oral MPA and cerebral meningioma. Associations for dMPA exposure were similar when using a non-active (OR 1.87, 95% CI 1.16-3.00) or active comparator (OR 1.87, 95% CI 0.98-3.56). These associations were strongest for prolonged exposure compared to a non-active (OR 5.40, 95% CI 2.71-10.76) and active comparator (OR 4.01, 95% CI 1.21-13.30).

**CONCLUSION:** The current results are consistent with the prior literature that dMPA exposure is associated with an increased likelihood of meningioma for prolonged use. More research is needed to examine whether the association is limited to a certain histology or grade of meningioma. Clinicians should consider discussing with patients these reported associations prior to using dMPA.

## BACKGROUND

Medroxyprogesterone acetate (MPA) is a synthetic progestin that is primarily pre-scribed for contraception and amenorrhea due to hormonal imbalances; however, it is also indicated in women who experience abnormal uterine bleeding to prevent endometrial hyperplasia, as palliative treatment in those with inoperable endometrial carcinomas, and in women experiencing pain from endometriosis [1, 2]. MPA binds to progesterone receptors in the hypothalamus, the female reproductive tract, pituitary gland which results in the inhibition of the release of gonadotropin-releasing hormone [1, 2]. Less frequent gonadotropin-releasing hormone release reduces the luteinizing hormone surge that occurs mid-cycle, preventing follicular maturation and ovulation [1]. Additionally, it can change the endometrium from proliferative to secretive, which makes implantation difficult.

MPA can be administered orally in tablet form at doses of 2.5,5, or 10 milligrams, subcutaneously at 104 mg/0.65 mL, or as an intramuscular injection at a 150 mg/mL dose [1]. The intramuscular injection version of MPA (known as depot medroxyprogesterone acetate [dMPA]) is administered once every three months and has a typical use failure rate of 4% [3]. Recent estimates from the United States suggest that 3% of women who reported using contraception between 2017 and 2019 used the MPA injection, while 25% of women aged 15-49 who have had intercourse reported using the MPA injection; however, these percentages varied based on certain sociodemographic characteristics [3, 4]. The MPA injection was more commonly used by young women, black women, lower-income and publicly insured women [3, 4]. Notably, four times as many women who were living be-low 100% of the federal poverty line reported using the injection compared to women living 400% above the poverty line, and the rate of usage among publicly insured patients was three times the rate of privately insured patients [3].

Progesterone has been shown in prior studies to play a role in the development and existence of meningiomas, which are a type of central nervous tumor [5-7]. Meningiomas are mostly benign and slow-growing tumors (i.e., World Health Organization grade 1) and are the most common type of brain tumor, accounting for 36% of all intracranial tumors [8]. They occur more frequently in females, and the WHO grade 1 tumors have a recurrence rate up to 25% [9]. The hormone dependency of meningiomas has been well studied. After puberty, three females develop meningiomas for every one male [10]. More-over, between 33 and 89% of meningiomas express progesterone receptors [5]; however, this percentage can vary depending on the pregnancy status of a woman and in what phase of the menstrual cycle a woman is [5, 10, 11]. All meningiomas that developed in pregnant or postpartum patients expressed progesterone receptors compared to 75.7% of meningiomas in non-pregnant female patients [5]. Additionally, increasing age was associated with decreasing expression of progesterone receptors in meningiomas according to a recent systematic review [5].

Research has identified an association between the use of progestogens and subsequent development of intracranial meningiomas [12-15]. Literature on the link between the intramuscular injection form of MPA and subsequent development of meningiomas is less common. Two prior population-based case control studies have identified an association between dMPA and intracranial meningiomas. The first, a matched case control study derived from the French National Health Data System Data-base, reported a near six-times higher odds of meningiomas compared to age and location matched controls [13]; however, the study was limited by low exposure to dMPA in this population with only 0.05% of cases and 0.01% of controls reporting use of dMPA [13]. The second study was a case control study conducted from a large private-insurance claims database based in the United States [14], reporting similar associations to the prior study. This case control study was based on employees of large companies who had private health insurance and could be generally healthier than a publicly insured population. Thus, the results may not generalize to a publicly insured patient population as several prior studies have noted differences in health care utilization and outcomes by type of health insurance in American patient populations [16-19]. In addition, both prior studies did not utilize an active comparator medication in their analyses, resulting in associations that may be biased due to confounding by factors related to the initiation of medication use.

To address these limitations, the objective of the current study was to assess whether the previously reported association between MPA and intracranial meningiomas is also observed in a population of publicly insured women in the United States while also advancing the research by utilizing an active comparator in addition to a non-active comparator.

## MATERIALS AND METHODS

### Data source

The current study utilized Alabama Medicaid administrative data from 2010 to 2023. The Medicaid claims data contains information on enrollment, demographic, inpatient and outpatient encounter claims, and pharmaceutical claims. The claims records, enrollment records, and demographics records were combined utilizing encrypted identification keys provided for both claims and persons enrolled.

### Study design and variables

Using a matched case-control design, cases were defined as females aged 18 to 55 years with an ICD-9 or ICD-10 diagnosis code for a cerebral meningioma that was benign (225.2, D32.0), malignant (192.1, C70.0), or of an unknown histological behavior (D42.0). For each case, the date earliest diagnosis was used as the case date. For each case, up to 10 female controls were matched on age±1 year and year of Medicaid enrollment; in addition, controls must have been enrolled for at least as long as the case.

For each case and matched controls, data was collected on ICD-9 and ICD-10 diagnoses for claims made within the three years prior to the case date and claimed medication dispensations at any point prior to the case date. For the former, Elixhauser comorbidities were determined for each study participant. The count of comorbidities was summed for each subject, and a categorical variable of unweighted Elixhauser comorbidity count was defined as zero or one comorbidity, two comorbidities, three comorbidities, or four or more comorbidities.

### Medication exposure

Exposure to MPA was determined separately for both oral MPA and dMPA. For both routes of administration, duration of exposure was determined, following the definition of Roland et al, as no exposure, short-term (i.e., exposure only for one year), and pro-longed-term exposure (i.e., exposure for one year followed by at least one dispensation the following year). There were 12 subjects (0.2% of the total population and all of whom were controls) that had exposure to both MPA and dMPA; these subjects were included in both the MPA and dMPA analyses. The same methodology was conducted for the active comparator exposure, defined as a claimed dispensation of norethindrone (alone or in combination) or of levonorgestrel in combination with another medication (e.g., levonorgestrel and ethinyl estradiol). Levonorgestrel alone is used an emergency contraceptive rather than continued contraceptive use and was excluded from consideration as an active comparator. These two medications were chosen due to their use as contraceptives and have not been reported to be associated with meningioma risk in cohort and case-control studies [20,21]. Using these variables, subjects were categorized as having no exposure (i.e., non-active comparator), exposure to an active comparator of levonorgestrel or norethindrone, and exposure to MPA (for the oral MPA analysis) or dMPA (for the injection MPA analysis).

### Statistical analysis

Age, race, and unweighted Elixhauser comorbidities were compared between case and matched controls utilizing a conditional logistic regression. Odds ratios (ORs) and associated 95% confidence intervals (CIs) for the association between MPA/dMPA expo-sure and cerebral meningioma were estimated from a conditional logistic regression. Models were adjusted for age, race, and categorical number of Elixhauser comorbidities, and separate models were created for any exposure, short-term exposure, and pro-longed-term exposure. In a sensitivity analysis, based on recommendations for dMPA not being used longer than two years [1], models were created after categorizing exposures (both MPA/dMPA and active comparator) more than one year prior to the case date and more than two years prior to the case date as non-exposures. SAS v9.4 was used for all analyses, and a two-tailed alpha of 0.05 was used for determination of statistical significance.

## RESULTS

The study population included 469 cases of meningiomas and 4,690 matched controls. A majority (90%) of patients identified as black or white, and there was no statistical difference of race between cases and controls (p=0.4874) (Table 1). Cases had a higher prevalence of four or more comorbidities compared to controls (63.5% vs 29.6%, p<0.0001). There was no difference between cases and controls in regard to comorbidities of AIDS (p=0.3829), alcohol abuse (p=0.1338), moderate or severe liver disease (p=0.1012), or pulmonary circulation disorders (p=0.2854). The prevalence of all other conditions comprising the Elixhauser comorbidity index were significantly different (p<0.05).

**Table 1.** Comparison of age and comorbidities of meningioma cases and matched controls.

|  | Cases (n=469) | Controls (n=4,690) | p-value |
| --- | --- | --- | --- |
| **Age** |  |  |  |
| 18-39 | 156 (33.3) | 1,562 (33.3) | 1.0000 |
| 40-55 | 312 (66.7) | 3,120 (66.7) |  |
| **Race** |  |  |  |
| Asian/Pacific Islander | 1 (0.2) | 13 (0.3) | 0.4874 |
| Black | 196 (41.8) | 2,059 (43.9) |  |
| White | 230 (49.0) | 2,100 (44.8) |  |
| Hispanic | 6 (1.3) | 76 (1.6) |  |
| American Indian/Alaskan Native | 3 (0.6) | 23 (0.5) |  |
| Other/Unknown | 33 (7.0) | 419 (8.9) |  |
| **Elixhauser comorbidity count** |  |  |  |
| 0-1 | 66 (14.1) | 2,476 (52.8) | <0.0001 |
| 2 | 47 (10.0) | 406 (8.7) |  |
| 3 | 58 (12.4) | 420 (8.9) |  |
| ≥4 | 298 (63.5) | 1,388 (29.6) |  |
| AIDS | 5 (1.1) | 33 (0.7) | 0.3829 |
| Alcohol abuse | 20 (4.3) | 141 (3.0) | 0.1338 |
| Deficiency anemias | 151 (32.2) | 778 (16.6) | <0.0001 |
| Autoimmune conditions | 41 (8.7) | 214 (4.6) | <0.0001 |
| Chronic blood loss anemia | 12 (2.6) | 59 (1.3) | 0.0232 |
| Leukemia | 5 (1.1) | 6 (0.1) | 0.0005 |
| Lymphoma | 4 (0.9) | 10 (0.2) | 0.0180 |
| Metastatic cancer | 24 (5.1) | 25 (0.5) | <0.0001 |
| Solid tumor without metastasis, in situ | 9 (1.9) | 42 (0.9) | 0.0369 |
| Solid tumor without metastasis, malignant | 90 (19.2) | 131 (2.8) | <0.0001 |
| Cerebrovascular disease | 83 (17.7) | 175 (3.7) | <0.0001 |
| Heart failure | 36 (7.7) | 230 (4.9) | 0.0091 |
| Coagulopathy | 21 (4.5) | 98 (2.1) | 0.0012 |
| Dementia | 17 (3.6) | 37 (0.8) | <0.0001 |
| Depression | 179 (38.2) | 1,066 (22.7) | <0.0001 |
| Diabetes with chronic complications | 74 (15.8) | 464 (9.9) | <0.0001 |
| Diabetes without chronic complications | 125 (26.7) | 749 (16.0) | <0.0001 |
| Drug abuse | 66 (14.1) | 416 (8.9) | 0.0002 |
| Complicated hypertension | 54 (11.5) | 293 (6.2) | <0.0001 |
| Uncomplicated hypertension | 274 (58.4) | 1,663 (35.5) | <0.0001 |
| Mild liver disease | 55 (11.7) | 259 (5.5) | <0.0001 |
| Moderate/Severe liver disease | 4 (0.9) | 16 (0.3) | 0.1012 |
| Chronic pulmonary disease | 142 (30.3) | 918 (19.6) | <0.0001 |
| Neurological disorders affecting movement | 31 (6.6) | 129 (2.8) | <0.0001 |
| Other neurologic disorders | 206 (43.9) | 155 (3.3) | <0.0001 |
| Seizures and epilepsy | 148 (31.6) | 302 (6.4) | <0.0001 |
| Obesity | 186 (39.7) | 1,021 (21.8) | <0.0001 |
| Paralysis | 26 (5.5) | 88 (1.9) | <0.0001 |
| Peripheral vascular disorders | 34 (7.2) | 200 (4.3) | 0.0027 |
| Psychoses | 90 (19.2) | 589 (12.6) | <0.0001 |
| Pulmonary circulation disorders | 10 (2.1) | 70 (1.5) | 0.2854 |
| Moderate renal failure | 32 (6.8) | 128 (2.7) | <0.0001 |
| Severe renal failure | 12 (2.6) | 56 (1.2) | 0.0156 |
| Hypothyroidism | 78 (16.6) | 417 (8.9) | <0.0001 |
| Other thyroid disease | 27 (5.8) | 171 (3.6) | 0.0232 |
| Peptic ulcer with bleeding | 32 (6.8) | 88 (1.9) | <0.0001 |
| Valvular disease | 39 (8.3) | 244 (5.2) | 0.0045 |
| Weight loss | 50 (10.7) | 261 (5.7) | <0.0001 |

\* Estimated from conditional logistic regression

### Non-active comparator

Oral exposure to MPA occurred for 3.2% for both cases and controls (Table 2). There was no association between any oral MPA exposure and subsequent development of cerebral meningiomas in both the crude (OR 1.01, 95% CI 0.59-1.75) and adjusted (OR 0.67, 95% CI 0.37-1.19) analyses. The lack of association remained for both short-term (adjusted OR 0.68, 95% CI 0.38-1.23) or prolonged-term (adjusted OR 0.48, 95% CI 0.06-3.70) expo-sures. Exposure to dMPA occurred among 6.0% of case and 3.6% of controls; in the adjusted model, cases were nearly twice as likely to have dMPA exposure than controls (OR 1.87, 95% CI 1.16-3.00). This association was only present for prolonged-term exposures (OR 5.40, 95% CI 2.71-10.76); a null effect was observed for short-term exposures (OR 0.96, 95% CI 0.52-1.78).

**Table 2.** Odds ratios* (ORs) and associated 95% confidence intervals (CIs) for the association between medroxyprogesterone acetate (MPA) exposure (compared to non-active comparator) and cerebral meningioma.

| **Exposure type** | **Cases**<br>**(n=469)** | **Controls**<br>**(n=4,690)** | **Crude OR**<br>**(95% CI)** | **Adjusted† OR**<br>**(95% CI)** |
| --- | --- | --- | --- | --- |
| **ORAL** |  |  |  |  |
| Non-active comparator (%) | 46 (93.0) | 4,375 (93.3) | Referent | Referent |
| MPA exposure (%) |  |  |  |  |
| Any | 15 (3.2) | 149 (3.2) | 1.01 (0.59-1.75) | 0.67 (0.37-1.19) |
| Short-term | 14 (3.1) | 132 (2.9) | 1.05 (0.60-1.85) | 0.68 (0.38-1.23) |
| Prolonged | 0 (0.0) | 14 (0.3) | 0.67 (0.09-5.06) | 0.48 (0.06-3.70) |
| **INJECTION** |  |  |  |  |
| Non-active comparator (%) | 421 (89.8) | 4,345 (92.6) | Referent | Referent |
| MPA exposure (%) |  |  |  |  |
| Any | 28 (6.0) | 170 (3.6) | 1.84 (1.18-2.87) | 1.87 (1.16-3.00) |
| Short-term | 14 (3.1) | 127 (2.8) | 1.12 (0.63-2.01) | 0.96 (0.52-1.78) |
| Prolonged | 14 (3.1) | 42 (0.9) | 3.51 (1.87-6.60) | 5.40 (2.71-10.76) |

\* Estimated from conditional logistic regression compared to a non-active comparator (i.e., no exposure to levonorgestrel or norethindrone)
† Adjusted for age, race, and number of Elixhauser comorbidities

### Active comparator

Similar to the non-active comparator analysis, there was no association between meningioma and oral MPA exposure overall (adjusted OR 0.73, 95% CI 0.34-1.55), in the short term (adjusted OR 0.75, 95% CI 0.34-1.67), or prolonged term (adjusted OR 0.39, 95% CI 0.04-3.74) (Table 3). No significant association was observed for dMPA exposure overall (adjusted OR, 95% CI 0.98-3.56); however, prolonged-term exposure to dMPA was associated with a four-fold increased odds of meningioma (adjusted OR 4.01, 95% CI 1.21-13.30).

**Table 3.** Odds ratios* (ORs) and associated 95% confidence intervals (CIs) for the association between medroxyprogesterone acetate (MPA) exposure (compared to active comparator) and cerebral meningioma.

|  | Cases<br>(n=469) | Controls<br>(n=4,690) | Crude OR<br>(95% CI) | Adjusted† OR<br>(95% CI) |
| --- | --- | --- | --- | --- |
| **ORAL MPA EXPOSURE** |  |  |  |  |
| Any (%) |  |  |  |  |
| MPA | 15 (3.2) | 149 (3.2) | 0.92 (0.45-1.91) | 0.73 (0.34-1.55) |
| Oral levonorgestrel/norethindrone | 18 (3.8) | 166 (3.5) | Referent | Referent |
| Short-term |  |  |  |  |
| MPA | 14 (3.0) | 134 (2.9) | 0.97 (0.45-2.10) | 0.75 (0.34-1.67) |
| Oral levonorgestrel/norethindrone | 15 (3.2) | 140 (3.0) | Referent | Referent |
| Prolonged |  |  |  |  |
| MPA | 1 (0.2) | 15 (0.3) | 0.43 (0.05-3.98) | 0.39 (0.04-3.74) |
| Oral levonorgestrel/norethindrone | 5 (1.1) | 32 (0.7) | Referent | Referent |
| **INJECTION MPA EXPOSURE** |  |  |  |  |
| Any (%) |  |  |  |  |
| MPA | 28 (6.0) | 170 (3.6) | 1.45 (0.78-2.69) | 1.87 (0.98-3.56) |
| Oral levonorgestrel/norethindrone | 20 (4.3) | 175 (3.7) | Referent | Referent |
| Short-term |  |  |  |  |
| MPA | 14 (3.0) | 127 (2.7) | 1.03 (0.48-2.21) | 1.13 (0.51-2.49) |
| Oral levonorgestrel/norethindrone | 16 (3.4) | 150 (3.2) | Referent | Referent |
| Prolonged |  |  |  |  |
| MPA | 14 (3.0) | 43 (0.9) | 2.01 (0.65-6.22) | 4.01 (1.21-13.30) |
| Oral levonorgestrel/norethindrone | 5 (1.1) | 30 (0.6) | Referent | Referent |

\* Estimated from conditional logistic regression
† Adjusted for age, race, and number of Elixhauser comorbidities

### Sensitivity analysis

Associations for oral MPA exposure were similarly decreased and not statistically significant when excluding exposures over a year prior to the case date and over two years prior to the case date (Table 4). For dMPA exposure, when excluding exposures a year after the case date, any dMPA exposure was associated with three-fold increased odds of meningioma in crude models (OR 3.31, 95% CI 1.73-6.32), an association that became stronger in adjusted models (OR 6.22, 95% CI 1.85-20.96). For prolonged exposure, an over six-fold increased odds of meningioma was observed in the adjusted model (OR 6.31, 95% CI 1.11-35.94). A similar pattern was observed after excluding exposures more than two years prior to the case date with significantly increased associations for both any dMPA exposure (OR 3.24, 95% CI 1.26-8.30) and prolonged exposure (OR 4.79, 95% CI 1.10-20.84).

**Table 4.** Odds ratios*† (ORs) and associated 95% confidence intervals (CIs) for the association between medroxyprogesterone acetate (MPA) exposure and cerebral meningioma excluding exposures one year and two years prior to case date

|  |  |  | Adjusted OR<br>(95% CI) |  |
| --- | --- | --- | --- | --- |
|  | Cases MPA<br>N (%) | Controls MPA<br>N (%) | vs non-active<br>comparator | vs active‡<br>comparator |
| **ORAL MPA EXPOSURE** |  |  |  |  |
| Excluding >1 year prior |  |  |  |  |
| Any | 3 (0.6) | 46 (1.0) | 0.35 (0.11-1.15) | 0.39 (0.11-1.43) |
| Short-term | 2 (0.4) | 37 (0.8) | 0.27 (0.06-1.17) | 0.30 (0.06-1.43) |
| Prolonged | 1 (0.2) | 9 (0.2) | 0.76 (0.09-6.15) | 0.74 (0.07-7.89) |
| Excluding >2 years prior |  |  |  |  |
| Any | 5 (1.1) | 66 (1.4) | 0.44 (0.17-1.12) | 0.49 (0.17-1.43) |
| Short-term | 4 (0.9) | 56 (1.2) | 0.41 (0.14-1.15) | 0.45 (0.14-1.47) |
| Prolonged | 1 (0.2) | 10 (0.2) | 0.65 (0.08-5.23) | 0.67 (0.06-7.03) |
| **INJECTION MPA EXPOSURE** |  |  |  |  |
| Excluding >1 year prior |  |  |  |  |
| Any | 15 (3.2) | 47 (1.0) | 3.31 (1.73-6.32) | 6.22 (1.85-20.96) |
| Short-term | 6 (1.3) | 29 (0.6) | 1.81 (0.70-4.70) | 5.30 (0.94-30.02) |
| Prolonged | 9 (1.9) | 18 (0.4) | 6.14 (2.53-14.90) | 6.31 (1.11-35.94) |
| Excluding >2 years prior |  |  |  |  |
| Any | 18 (3.8) | 76 (1.6) | 2.73 (1.52-4.91) | 3.24 (1.26-8.30) |
| Short-term | 8 (1.7) | 49 (1.0) | 1.48 (0.65-3.35) | 2.18 (0.63-7.59) |
| Prolonged | 10 (2.1) | 27 (0.6) | 5.71 (2.51-13.00) | 4.79 (1.10-20.84) |

\* Estimated from conditional logistic regression
† Adjusted for age, race, and number of Elixhauser comorbidities
‡ Dispensation of norethindrone (alone or in combination) or levonorgestrel in combination (e.g., levonorgestrel and ethinyl estradiol)

## DISCUSSION

The results of this matched case-control study showed a strong and persistent association between dMPA exposure and development of cerebral meningiomas in a publicly insured patient population in the United States. This association was limited to exposures for at least two continuous years, and, in sensitivity analyses, limiting exposures to those recent to the case date increased the strength of associations. No significant associations were observed for oral MPA exposure.

The associations reported in the current study are similar to associations previously reported [13, 14, 23] and support the findings of increased likelihood of meningioma for use of MPA in a disproportionality study utilizing data from the Food and Drug Administration’s Adverse Events Reporting System [24]. In a case-control study based on the National Health Data System in France [13], though there was low overall dMPA expo-sure (0.05% of cases, 0.01% of controls), prolonged dMPA exposure (using the same definition as in the current study) was associated with 5.6 times the odds of developing meningioma compared to the controls, similar to the 5.4 association for dMPA in the current study. Short term exposure could not be analyzed due to only two participants having short-term exposure to dMPA; however, the results of the current study suggest no association is present within the first year of use. Another case control study conducted from a large private-insurance claims database based in the United States [14] reported that an exposure of two or more years to the dMPA was associated with over twice the odds of developing a cerebral meningioma compared to the controls even after accounting for age and comorbidities [14]. The association for one year of use was similar (OR 1.35) to what the current study reports. Differences in the association with the current study are likely due to differences in the study population, with the current study including only those with public insurance and the previous study including only those with commercial, employer-provided insurance.

Other studies have reported meningioma occurrence among those with long-term dMPA use. In a retrospective study among women receiving treatment for meningiomas at the University of Pittsburgh Medical center between 2014 and 2021 [22], twenty-five women had a total of 49 meningiomas which all stained positively in progesterone immunohistochemical staining. The average time of dMPA use was 15.5 years among the 25 women. Additionally, of the ten women instructed to stop using dMPA, there was evident tumor shrinkage in half of them, further supporting evidence of a potential link between dMPA usage and development of cerebral meningiomas [22]. Two additional studies in Indonesia and Sweden looked at various types of contraceptive use and development of meningiomas or gliomas [15, 24]. A case-control study in Indonesia found elevated odds of meningiomas with ten or more years of hormonal contraceptive use compared to those with no use (OR: 18.22); however, dMPA was combined with other types of contraception, and the isolated effect of dMPA cannot be ascertained [24]. The Sweden-based study found that women who had used long-acting hormonal contraceptives (subdermal implants, injections, hormonal intrauterine devices) for between five and nine years had 2.5 times the odds of meningioma, but there was no association with development of gliomas [15].

The persistence of a strong association in different populations between prolonged dMPA use and later development of cerebral meningiomas supports the notion that there may be a causal link between exposure to dMPA and meningiomas. Moreover, there is a biological explanation for the observed associations between dMPA use and meningiomas. A recent systematic review on hormone reception expression in meningiomas noted that slightly over two-thirds of meningiomas expressed progesterone receptors, but this percentage varied depending on the hormone treatment and pregnancy status of the women [5]. Progesterone levels are elevated during pregnancy, and all of the meningiomas that developed during pregnancy expressed progesterone receptors [5]. Further, MPA has one of the highest progesterone receptor affinities of the progestins [25]. Progesterone receptor status of meningiomas has been associated with location (with a higher prevalence in skull-base meningiomas [26,27]), recurrence [27], and WHO grade [27,28]. For the latter, higher progesterone receptor prevalence is associated with lower grade (WHO grade 1) meningiomas. These grade 1 meningiomas are benign, but treatment, when including surgical resection, can be costly (with a reported median cost of approximately $40,000 for the initial admission [29]) and has a reported complication rate as high as 41% with the most common complications including seizures, dysrhythmia, intracranial hemorrhage, and cerebral artery occlusion [29].

### Limitations

The results of the current study need to be viewed in light of its limitations. First, the study was based on data from medical and pharmaceutical claims of publicly insured women in the U.S., which has two limitations. First, it is only known that a Medicaid beneficiary received a prescription for oral MPA; thus, it is possible that exposure misclassification occurred for oral MPA if the enrollee did not use the medication. This misclassification is not present for dMPA as it is a one-time injection. Second, results may not be generalizable to all women. As another limitation, while the associations in this study were adjusted for age, race and comorbidities, residual confounding may still be present. Another limitation is that this study could not consider grade or pathology of the meningiomas, therefore, it could not examine whether there is a link between prolonged dMPA use and the various grades of meningiomas. Approximately 75% of meningiomas are benign and slow-growing tumors (i.e. grade 1); however, even benign tumors can cause significant debilitating neurologic complications which warrants further investigation. Finally, there was a large discrepancy in prevalence of comorbidities between cases and controls; however, when limiting logistic models to those with three or more comorbidities and, separately, 0-2 comorbidities, associations were similar to reported associations.

## CONCLUSIONS

The current study noted elevated odds of cerebral meningiomas with at least two years of exposure to dMPA, which is consistent with prior studies [13-15, 22, 24] and pro-vides further evidence of the link between dMPA exposure and development of meningiomas. In particular, the strong association observed in the three previous case-control studies and the current study, the observation in the current and prior studies finding the association only among dMPA rather than oral MPA, and the progesterone receptor ex-pression in meningiomas provides evidence of a potential causal link between dMPA use and subsequent development of cerebral meningioma.

Clinicians should consider, for patients who are considering dMPA as their contraceptive method, informing the patients of the risks of developing cerebral meningiomas. Future research needs to investigate whether dMPA exposure is associated with specific tumor grades and pathologies.

## Data Availability

Data are available from request to the Centers for Medicare and Medicaid Services

https://resdac.org/

## REFERENCES

1. Sathe A, Patel P, Gerriets V. Medroxyprogesterone. [Updated 2024 Feb 29]. In: StatPearls [Internet]. Treasure Island (FL): StatPearls Publishing; 2024 Jan-. Available from: https://www.ncbi.nlm.nih.gov/books/NBK559192/ Accessed 6/5/2025.

2. Edwards M.; Can A.S. Progestins. In StatPearls; StatPearls Publishing: Treasure Island (FL):StatPearls Publishing; 2024 Jan. Available from: https://www.ncbi.nlm.nih.gov/books/NBK563211/. Accessed 6/5/2025.

3. DMPA Contraception Injection: Use and Coverage.; Available from: https://www.kff.org/womens-health-policy/fact-sheet/dmpa-contraceptive-injection-use-and-coverage/. Access 6/5/2025.

4. Haakenstad, A.,, Angelino, O.,, Irvine, C.M.S.,, Bhutta, Z.A.,, Bienhoff, K.,, Bintz, C.,, Causey, K.,, Dirac, M.A.,, Fullman, N.,, Gakidou, E.,, et al. Measuring Contraceptive Method Mix, Prevalence, and Demand Satisfied by Age and Marital Status in 204 Countries and Territories, 1970-2019: A Systematic Analysis for the Global Burden of Disease Study 2019. Lancet 2022, 400, 295–327, doi:10.1016/S0140-6736(22)00936-9.

5. Agopiantz, M.,, Carnot, M.,, Denis, C.,, Martin, E.,, Gauchotte, G. Hormone Receptor Expression in Meningiomas: A Systematic Review. Cancers (Basel) 2023, 15, 980, doi:10.3390/cancers15030980.

6. Guevara, P.,, Escobar-Arriaga, E.,, Saavedra-Perez, D.,, Martinez-Rumayor, A.,, Flores-Estrada, D.,, Rembao, D.,, Calderon, A.,, Sotelo, J.,, Arrieta, O. Angiogenesis and Expression of Estrogen and Progesterone Receptors as Predictive Factors for Recurrence of Meningioma. J Neurooncol 2010, 98, 379–384, doi:10.1007/s11060-009-0086-z.

7. Kuroi, Y.,, Matsumoto, K.,, Shibuya, M.,, Kasuya, H. Progesterone Receptor Is Responsible for Benign Biology of Skull Base Meningioma. World Neurosurg 2018, 118, e918– e924, doi:10.1016/j.wneu.2018.07.100.

8. Ostrom, Q.T.,, Price, M.,, Neff, C.,, Cioffi, G.,, Waite, K.A.,, Kruchko, C.,, Barnholtz-Sloan, J.S. CBTRUS Statistical Report: Primary Brain and Other Central Nervous System Tumors Diagnosed in the United States in 2015-2019. Neuro Oncol 2022, 24, v1–v95, doi:10.1093/neuonc/noac202.

9. Louis, D.N.,, Perry, A.,, Reifenberger, G.,, von Deimling, A.,, Figarella-Branger, D.,, Cavenee, W.K.,, Ohgaki, H.,, Wiestler, O.D.,, Kleihues, P.,, Ellison, D.W. The 2016 World Health Organization Classification of Tumors of the Central Nervous System: A Summary. Acta Neuropathol 2016, 131, 803–820, doi:10.1007/s00401-016-1545-1.

10. Claus, E.B.,, Bondy, M.L.,, Schildkraut, J.M.,, Wiemels, J.L.,, Wrensch, M.,, Black, P.M. Epidemiology of Intracranial Meningioma. Neurosurgery 2005, 57, 1088–1095; discussion 1088-1095, doi:10.1227/01.neu.0000188281.91351.b9.

11. Hatiboglu, M.A.,, Cosar, M.,, Iplikcioglu, A.C.,, Ozcan, D. Sex Steroid and Epidermal Growth Factor Profile of Giant Menin-giomas Associated with Pregnancy. Surg Neurol 2008, 69, 356–362; discussion 362-363, doi:10.1016/j.surneu.2007.03.013.

12. Hoisnard, L.,, Laanani, M.,, Passeri, T.,, Duranteau, L.,, Coste, J.,, Zureik, M.,, Froelich, S.,, Weill, A. Risk of Intracranial Meningioma with Three Potent Progestogens: A Population-Based Case-Control Study. Eur J Neurol 2022, 29, 2801–2809, doi:10.1111/ene.15423.

13. Roland, N.,, Neumann, A.,, Hoisnard, L.,, Duranteau, L.,, Froelich, S.,, Zureik, M.,, Weill, A. Use of Progestogens and the Risk of Intracranial Meningioma: National Case-Control Study. BMJ 2024, 384, e078078, doi:10.1136/bmj-2023-078078.

14. Griffin, R.L. The Association between Medroxyprogesterone Acetate Exposure and Meningioma. Cancers (Basel) 2024, 16, 3362, doi:10.3390/cancers16193362.

15. Wigertz, A.,, Lönn, S.,, Mathiesen, T.,, Ahlbom, A.,, Hall, P.,, Feychting, M.,, Swedish Interphone Study Group Risk of Brain Tumors Associated with Exposure to Exogenous Female Sex Hormones. Am J Epidemiol 2006, 164, 629–636, doi:10.1093/aje/kwj254.

16. Freeman, J.D.,, Kadiyala, S.,, Bell, J.F.,, Martin, D.P. The Causal Effect of Health Insurance on Utilization and Outcomes in Adults: A Systematic Review of US Studies. Med Care 2008, 46, 1023–1032, doi:10.1097/MLR.0b013e318185c913.

17. Hoffman, C.,, Paradise, J. Health Insurance and Access to Health Care in the United States. Ann N Y Acad Sci 2008, 1136, 149–160, doi:10.1196/annals.1425.007.

18. Zhao, J.,, Han, X.,, Nogueira, L.,, Fedewa, S.A.,, Jemal, A.,, Halpern, M.T.,, Yabroff, K.R. Health Insurance Status and Cancer Stage at Diagnosis and Survival in the United States. CA Cancer J Clin 2022, 72, 542–560, doi:10.3322/caac.21732.

19. Jain, V.,, Venigalla, S.,, Sebro, R.A.,, Karakousis, G.C.,, Wilson, R.J.,, Weber, K.L.,, Shabason, J.E. Association of Health Insurance Status with Presentation, Treatment and Outcomes in Soft Tissue Sarcoma. Cancer Med 2019, 8, 6295–6304, doi:10.1002/cam4.2441.

20. Korhonen, K.,, Auvinen, A.,, Lyytinen, H.,, Ylikorkala, O.,, Pukkala, E. A Nationwide Cohort Study on the Incidence of Meningioma in Women Using Postmenopausal Hormone Therapy in Finland. Am J Epidemiol 2012, 175, 309–314, doi:10.1093/aje/kwr335.

21. Roland, N.,, Kolla, E.,, Baricault, B.,, Dayani, P.,, Duranteau, L.,, Froelich, S.,, Zureik, M.,, Weill, A. Oral Contraceptives with Progestogens Desogestrel or Levonorgestrel and Risk of Intracranial Meningioma: National Case-Control Study. BMJ 2025, 389, e083981, doi:10.1136/bmj-2024-083981.

22. Abou-Al-Shaar, H.,, Wrigley, R.,, Patel, A.,, Mallela, A.N.,, Zenonos, G.A.,, Gardner, P.A. Skull Base Meningiomas as Part of a Novel Meningioma Syndrome Associated with Chronic Depot Medroxyprogesterone Acetate Use. In Proceedings of the Journal of Neurological Surgery Part B: Skull Base; Georg Thieme Verlag KG, February 1 2023; Vol. 84, p. S231.

23. Frey, C.,, Etminan, M. Disproportionality Analysis of Progestogens and Estrogens Demonstrates Increased Meningioma Risk. J Clin Neurosci 2025, 137, 111328, doi:10.1016/j.jocn.2025.111328.

24. Wahyuhadi, J.,, Heryani, D.,, Basuki, H. Risk of Meningioma Associated with Exposure of Hormonal Contraception. A Case Control Study. Majalah Obstetri & Ginekologi 2018, 26, 36– 41, doi:10.20473/mog.V26I12018.36-41.

25. Pletzer, B.,, Winkler-Crepaz, K.,, Maria Hillerer, K. Progesterone and Contraceptive Progestin Actions on the Brain: A Systematic Review of Animal Studies and Comparison to Human Neuroimaging Studies. Front Neuroendocrinol 2023, 69, 101060, doi:10.1016/j.yfrne.2023.101060.

26. Maiuri, F.,, Mariniello, G.,, Guadagno, E.,, Barbato, M.,, Corvino, S.,, Del Basso De Caro, M. WHO Grade, Proliferation Index, and Progesterone Receptor Expression Are Different According to the Location of Meningioma. Acta Neurochir (Wien) 2019, 161, 2553–2561, doi:10.1007/s00701-019-04084-z.

27. Maiuri, F.,, Mariniello, G.,, de Divitiis, O.,, Esposito, F.,, Guadagno, E.,, Teodonno, G.,, Barbato, M.,, Del Basso De Caro, M. Progesterone Receptor Expression in Meningiomas: Pathological and Prognostic Implications. Front Oncol 2021, 11, 611218, doi:10.3389/fonc.2021.611218.

28. Roser, F.,, Nakamura, M.,, Bellinzona, M.,, Rosahl, S.K.,, Ostertag, H.,, Samii, M. The Prognostic Value of Progesterone Receptor Status in Meningiomas. J Clin Pathol 2004, 57, 1033–1037, doi:10.1136/jcp.2004.018333.

29. Connolly, I.D.,, Cole, T.,, Veeravagu, A.,, Popat, R.,, Ratliff, J.,, Li, G. Craniotomy for Resection of Meningioma: An Age-Stratified Analysis of the MarketScan Longitudinal Database. World Neurosurg 2015, 84, 1864–1870, doi:10.1016/j.wneu.2015.08.018.

